# “I had made the decision, and no one was going to stop me” —Facilitators of PrEP adherence during pregnancy and postpartum in Cape Town, South Africa

**DOI:** 10.1101/2020.11.23.20236729

**Authors:** Dvora L. Joseph Davey, Lucia Knight, Jackie Markt-Maloney, Nokwazi Tsawe, Yolanda Gomba, Nyiko Mashele, Kathryn Dovel, Pamina Gorbach, Linda-Gail Bekker, Thomas J. Coates, Landon Myer

## Abstract

**Introduction:** HIV incidence is high during pregnancy and postpartum in many settings. PrEP is safe and effective but requires adherence during potential HIV exposure, yet the facilitators of high maternal adherence are not well understood in high HIV burden settings.

**Methods:** We conducted semi-structured interviews with women who reported high adherence (PrEP use ≥ 25 days in last 30-days) within a PrEP service for pregnant and postpartum women located in a large primary care facility in a high-HIV burden township. Topics for interviews included: individual/interpersonal risk, disclosure, anticipated PrEP stigma, safety, side-effects, and facility-level factors effecting adherence. A thematic approach guided an iterative process of coding (reviewed to ensure intercoder reliability) and analysis using NVivo 12.

**Results:** We interviewed 25 postpartum women with high PrEP adherence who were on PrEP for a median of 9-months, median age 26-years, and median baseline gestational age 24-weeks. Themes identified as key drivers of optimal PrEP use were HIV risk perception – primarily due to partner’s perceived risky sexual behaviors and unknown serosatus—and a strong desire to have a baby free of HIV. Reported disclosure of PrEP use to family, partners and friends facilitated PrEP adherence. Women continued PrEP postpartum because they felt empowered by PrEP and did not want to “go backwards” and increase their HIV risk as before PrEP. Women who reported high adherence all discussed having community support and reminders to take PrEP on time. The primary barriers were anticipated or experienced stigma, which most overcame through education of partners/family about PrEP. Pregnant women experienced transient side effects, but found ways to continue, including taking PrEP at night. Women believed PrEP education and counselling were accessible when integrated into antenatal care which contributed to continued PrEP use.

**Conclusions:** Facilitators of optimal PrEP use through pregnancy and postpartum included fear of HIV acquisition for self and infant, mostly due to partner sexual behaviors and unknown serostatus, along with PrEP disclosure, and encouragement from partners and family. PrEP programs for pregnant and postpartum women should integrate strategies to assist women with realistic appraisals of risk and teach skills for securing support for significant others.

## Introduction

Women in sub-Saharan Africa face a high risk of HIV acquisition during pregnancy and breastfeeding.[2] While services to eliminate mother-to-child HIV transmission (EMTCT) have expanded rapidly in the region, few primary prevention interventions exist for pregnant women who initially test HIV-negative. This is a missed opportunity with implications for the woman, but also her partner and infant.[3, 4] Acute HIV infection in pregnancy and postpartum increases the risk of vertical transmission, accounting for an estimated third of vertical HIV transmissions.[5, 6] To protect women and contribute to EMTCT, the World Health Organization (WHO) recommends offering pre-exposure prophylaxis (PrEP) to pregnant and postpartum women at risk of HIV acquisition.[7-9] In South Africa, an estimated 76,000 infant HIV cases are expected between 2020-2030, though this could reduce significantly if pregnant women initiate and persist on PrEP.[10]

A recent systematic review provides no safety-related rationale for prohibiting PrEP during pregnancy and/or breastfeeding.[11] While safety data are reassuring, recent pharmacokinetic studies reveal that tenofovir diphosphate concentrations were one-third lower in pregnancy compared to postpartum women, highlighting how essential daily PrEP use is in pregnant women.[12, 13] There are numerous barriers to optimal PrEP use in pregnant and postpartum women across health facility-, interpersonal and intrapersonal-levels. [11, 14, 15] [16]

Within a larger study of PrEP in pregnancy, we conducted in-depth interviews with postpartum women who started PrEP in pregnancy and reported high levels of adherence though the transition to postpartum. The aim of this study is to understand the positive drivers of PrEP initiation and adherence in pregnant and postpartum women to inform future interventions to optimize PrEP use in this vulnerable population.

## Methods

The PrEP in pregnancy and postpartum (PrEP-PP) study is a prospective cohort enrolling adolescent girls and women at one primary care, public health clinic in Cape Town, South Africa (NCT03902418). Participants are enrolled at their first antenatal care (ANC) visit and followed participants through 12-months postpartum. Inclusion criteria include being: 1) =>16 years old, 2) confirmed HIV-negative 3) intending to stay in Cape Town, 4) confirmed pregnancy status, and 5) absence of medical contraindications to PrEP. Following the baseline survey, women are counselled about the risk of HIV in pregnancy and postpartum, including the benefits and risks of taking PrEP. If women wish to start PrEP, nurses prescribe a one-month supply of Truvada® (tenofovir disoproxil fumarate/emtricitabine). Participants on PrEP receive an invitation card to return for HIV testing and counselling, refill prescriptions and adherence counselling.

For this qualitative sub-study, we purposively recruited for homogeneity 25 postpartum between July and September, 2020. Enrolment was from the subset of women in the PrEP-PP cohort who were: 1) in study for 3+ months, 2) delivered a live infant, 3) reported high PrEP adherence in pregnancy and postpartum (defined as =>25 of last 30 days on PrEP), and attended all PrEP collection visits. Consenting participants received 120 Rand (approximately $8 USD) in grocery vouchers for their time and transport costs at each study visit.

One trained, female, isiXhosa speaking interviewer conducted a semi-structured interview in the local language (isiXhosa). Participants were interviewed telephonically due to the COVID-19 lockdown and associated risk (n=19 interviews), or in a private room within the health facility (n=6 interviews; both participant and interviewer wore masks and sat >1.5 meters apart). Interviews lasted approximately 30-40 minutes. Interviews were audio-recorded, translated into English, and transcribed by two independent trained research staff. The Interviewer reviewed all transcriptions and translations for accuracy and made necessary corrections.

A thematic approach guided an iterative process of coding and data analysis in NVivo 12 (QSR International, Victoria, Australia).[17] The analysis integrated memo writing and the concept mapping to refine understandings of PrEP adherence and develop themes that facilitated adherence. Three trained research assistants (YG, JMM, NT) conducted the analysis led by the study PI. The Consolidated Criteria for Reporting Qualitative Research (COREQ) checklist guided the analysis and reporting.[18]

### Conceptual framework

We used an adapted version of Ickovics’ and Meisler’s conceptual framework on factors affecting adherence in HIV treatment [1] that was used [19, 20] to situate and organise the findings into the broader context of the intersection between individual women and biomedical interventions **(Figure 1)**. The conceptual framework considered: 1) individual, 2) HIV factors, 3) pill safety/side-effects, and 4) facility-level factors explored from the participants’ perspective. The interview guide was developed to explore these factors in more depth.

**Figure 1.**
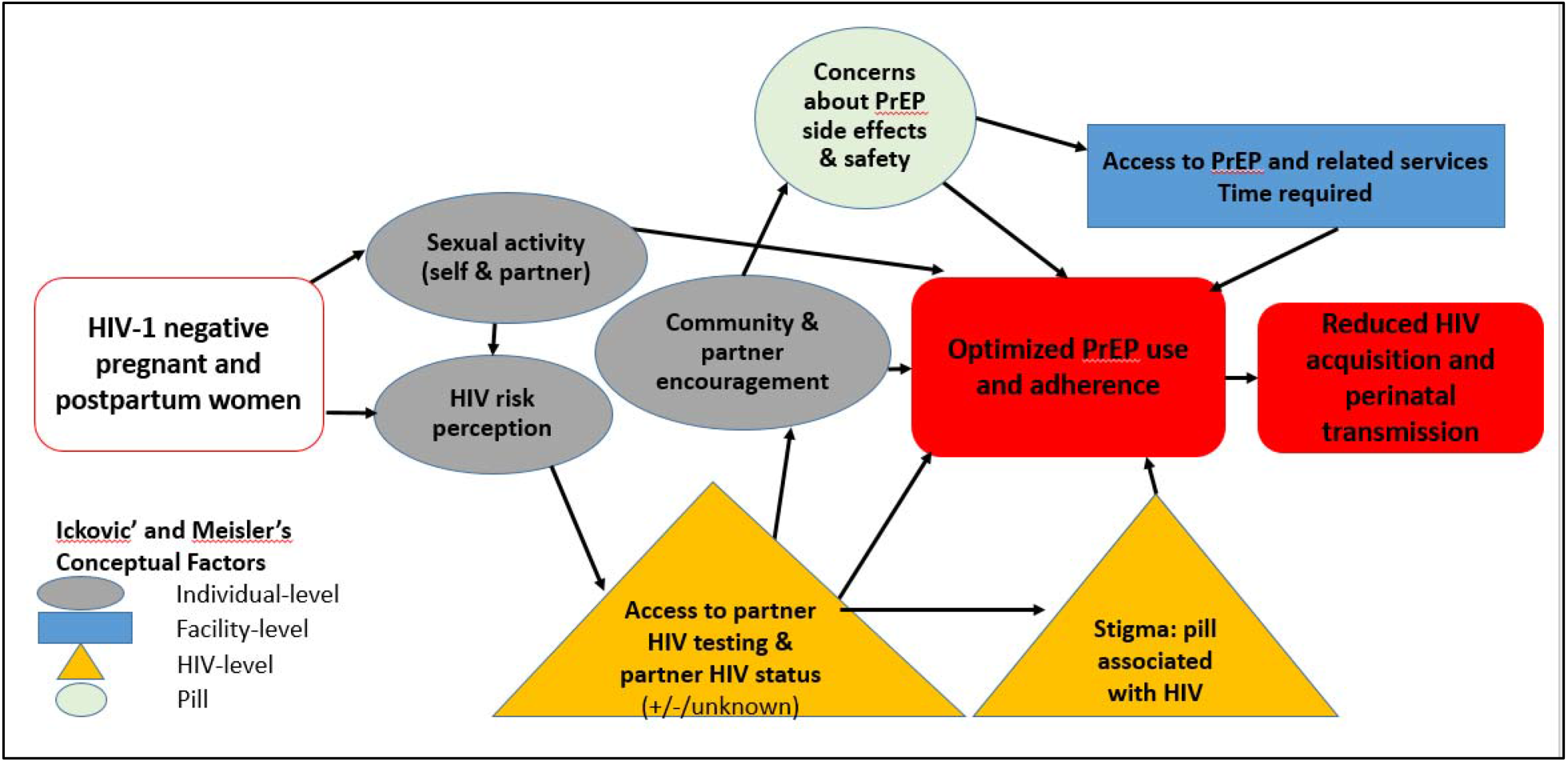
PrEP-PP conceptual model of individual, disease, facility level and pill factors associated with PrEP adherence adapted from Ickovics & Meisler [1]

### Ethics

The study was approved by the Human Research Ethics Committee at the University of Cape Town (#297/2018) and the University of California, Los Angeles Institutional Review Board (IRB#18-001622). All participants provided written informed consent during study enrolment.

## Results

### Participant characteristics

Participants were 16 to 43 years old and had been on PrEP for a median of 9 months. Almost all participants reported vaginal sex during pregnancy, fewer than one-third were married or cohabiting with a partner, and several reported multiple sex partners in the past year.

#### Individual factors

Women reported various reasons for why they initiated PrEP in pregnancy, as well as why they persisted on PrEP during pregnancy and the postpartum period. Most women decided to start PrEP to protect their baby from HIV. After taking PrEP for months, women reported not wanting to “go backwards” and being at risk of acquiring HIV again. One woman said, “I had made the decision, and no one was going to stop me. I was not going to go backwards*”* (PID005, 24 years). Others reported a feeling of relief or happiness while taking PrEP because they felt protected against HIV and “safe” during pregnancy. Below we present specific factors that encouraged women to start and stay on PrEP.

Women reported a period of postpartum abstinence ranging from weeks to months after giving birth. Despite this, the enrolled women who reported high adherence did not report stopping PrEP in this time. As one woman noted: “It [taking PrEP] was routine. I was not forgetting to take it anymore. Stopping or taking a break never came to mind.” (PID005, 24 years) Several women mentioned having more than one sex partner during pregnancy and postpartum and that PrEP helped them feel safer, especially with condomless sex.

### Motivation of HIV-free infant

A desire to remain HIV uninfected and have an HIV-free infant were strong motivators to continue PrEP.

> *It [PrEP] is protecting the unborn baby. It was very important that I protect the baby. I knew I was negative, so I didn’t know what the situation would be when the baby came. So, I was interested [in PrEP] because I wanted nothing [to happen] as I can see the conditions of people who are infected. What was important was protection, then I made the decision*. (PID67, 41 years)

This focus on protecting the baby was clearly presented and reinforced by the project staff during enrolment and influenced women’s thinking, for example: “They [project staff] explained that when you are breastfeeding, it is important to take these pills because they protect the baby. I plan on continuing because I am still breastfeeding. (PID152, 34 years)

Another mother took PrEP because her child was still too young for her to get infected and she wanted to be there for the child. A few women also credited PrEP for their babies being born HIV negative. “The baby was fine when I had her. She didn’t have a problem. I trust that it was the pills I was taking.” (PID152, 34 years)

Overall, women who adhered to PrEP expressed high satisfaction with PrEP feeling empowered to prevent HIV in their infants also contributing to their confidence in continuing use.

### Partner HIV risk and HIV testing

Women’s fear of HIV acquisition among their child, and their own seroconversion, was frequently linked to women’s perceived risk resulting from their partner’s risky sexual behaviour.

> *Even if you sleep with your partner without protection [condoms] or if you don’t know what he is up to, if you use PrEP, you know you are safe. That is the main reason that made me take PrEP … He loves girls; He is not trustworthy*. (PID156, 26 years)
>
> Fear of partners’ risky sexual behaviour was exacerbated by being pregnant and resulting limited willingness to have sex during this time.

> *When you are pregnant, you don’t get to be with your partner a lot especially if you don’t live together. So you don’t know what he is doing and with whom*. (PID 293, 20 years)
>
> Known or perceived risky sexual behavior by their partners lead some women to believe their partner was likely HIV-infected. However, most women reported not knowing their partner’s HIV status or his refusal to get tested. For example, “I took PrEP because of my partner’s behavior. Based on how he behaves he can even bring disease to me.*”* (PID114, 19 years)

Other women described forcing their partners to test, or not trusting them if they reported prior testing.

> *The people we live with nowadays are unfaithful. You would ask a person to go for HIV testing, they would lie and say they have but to find out that they didn’t even go for testing. So, this (taking PrEP) is my way of protecting myself*. (PID120, 32 years)
>
> Other women stated they had tested with their partners in the past, but that their status may have changed due to their sexual behavior. Partner testing was infrequent and untrusted by women, especially those who knew their partners had other sex partners.

In addition, few women reported using condoms prior to PrEP start, or during PrEP use, in many cases because of their partner’s choice.

> *He threw that condom away, because he was saying he will not use a condom. I told him this (condom use) was in the interest of our baby. Initially he thought I was cheating*. (PID120, 32 years)

As a result, PrEP helped the women feel protected or safe when they had sex with their partners who they thought were risky partners, or when navigating condom use was difficult. Women’s newfound sense of safety empowered them to continue to take PrEP. For example: “Now, I feel comfortable even when I don’t use a condom. I tell myself that I am alright.” (PID195, 25 years)

### Community PrEP encouragement

Encouragement from their primary partner, a friend or relative regarding their decision to start or continue taking PrEP was an important driver of consistent PrEP use.

> *She (mother) liked it and [said] that I should continue. Another person who knows is the father of my child, he knows that I am using PrEP. He didn’t have a problem with it and said I should continue. I had made the decision, and no one was going to stop me*. (PID067, 41 years)

Some were even interested in beginning PrEP themselves: “He asked how he could get it…..He even said that they should be available for everyone and not just pregnant people…” (PID195, 25 years)

A few women expressed concern about anticipated conflict or discouragement but were pleasantly surprised by their partners’ responses.

> *I was concerned that my partner will tell me about the child. [Him questioning me about] taking a pill during pregnancy and how can I trust them [pills]. But I decided to be courageous and tell him because he will find out, he will see it on the [clinic] card. Luckily, it was received by someone who understood it. I told him about it the way I was told…He was not stubborn and said I should continue*. (PID067, 41 years).

Others stated that their partner was concerned about the safety of taking PrEP during pregnancy and required additional information about PrEP to accept or support them:

> *…*.*He didn’t understand what PrEP even when I explained it. He said I was taking pills that would endanger his baby, so we had a fight. He threw the pills in the bin. I spoke to them [PrEP study staff] and they advised me on what I should do*.
>
> *They gave me a pamphlet and I sat him down and explained how it works. Only then did he believe that it won’t endanger the child*. (PID148, 35 years)

Several partners even helped remind women to take PrEP, this type of practical support may facilitate adherence: “sometimes he would remind me to take my tablets even before nine…. I won’t lie he encourages me!” (PID147, 42 years)

Many participants received encouragement from relatives, particularly immediate family members like parents and siblings.

> *…My sister was happy for me. She said it was the first time she was hearing about it, but she said it sounded interesting when I explained it…*.*Luckily, she had not heard about it and liked it, she said I should continue*. (PID067, 41 years)

Others said they had personal agency, regardless of what their community said. Partner, family and friend encouragement following disclosure about taking PrEP was a significant driver in PrEP adherence.

#### HIV-related factors

### Stigma related to taking PrEP

Several women reported concerns about disclosing PrEP use because they feared, or had experienced, community stigmatization. One woman noted her fear of being assumed to be living with HIV: “I was concerned that they would say it is not PrEP….Saying that I am hiding, that [the pills] are something else [ART], that they are not for what I say they are for.” (PID148, 35 years)

Other women supported this by reporting that on disclosing their PrEP use they were questioned about their HIV status. “They (friends) listened. They opened them….They looked and asked why they look like ART. I said I was not told that they are ART.*”* (PID178, 25 years)

Many women overcame that stigma by being open about their use of PrEP, and educating others about the benefits of taking PrEP. Despite some fear and reports of stigma, no participants reported that this affected their adherence. Most reported that in the end they were taking PrEP for themselves, not others.

### Concerns about PrEP safety and experiences of side effects

Pregnant women initiated PrEP at their first antenatal visit, at about 19 week’s gestation though many started earlier. Most women reported experiencing mild side effects, or morning sickness, including vomiting, nausea, diarrhoea and headaches. Some did not have any symptoms, but were concerned about getting side effects when they started PrEP.

> *It does take 2-3 days with any pill, having those side effects. Luckily, for me it only took one day. It (counselling) made me want to try it out myself and not hear about it from others*. (PID67, 41 years)
>
> Several women did experience side effects, when they started PrEP, and fear of future side effects motivated them to continue taking.

> *When I started, I was vomiting so I thought I would stop taking it, but I thought, what will I do if I stop. I changed, I was taking it in the morning, I changed and took it at night, then I was fine*. (PID43, 36 years)

Other participants were concerned about the safety of PrEP during pregnancy and breastfeeding, but were counselled on the risks and benefits. While women were concerned about, or experienced, transient side effects many continued taking PrEP because they did not want to go back to having side effects again. Women who reported high adherence were able to manage side effects and get over the transient side effects.

#### Facility-level factors

Participants learned about PrEP during the study and appreciated that education was integrated into ANC. In addition, participants appreciated the detailed counseling they received as part of the project.

> *They [project staff] also taught us about side effects that can manifest to some of us since our bodies are not the same. The explanation we get here is different from that we get in the clinics…they explained everything and that encouraged me to continue (on PrEP)*. (PID006, 16 years)

The counseling was also a key motivator for starting and staying on PrEP.

> *She (study counselor) said that when I use PrEP…. But what happened is, when I started using PrEP, I had those things, feeling like I will vomit. But because I was told about them, I knew I was not sick, it was just the pills I was taking*. (PID067, 41years)

All participants recommended continuing the offer of PrEP in antenatal care. Some women made suggestions for other pregnant women: “I would tell her to take PrEP, because it has worked for me and that I have also started it while I was pregnant. Even my child was okay, I gave birth to a healthy child.” (PID333, 23 years)

Participants liked that the offer of PrEP counseling and services were integrated into their antenatal and postnatal care services, and did not feel that the counseling or time spent was a burden on initiating or persisting on PrEP.

## Discussion

This study provides novel evidence on the facilitators of consistent PrEP use in pregnancy and postpartum. Individual level risk and risk perception promoted women to start and stay on PrEP, especially perceived HIV risk related to partners’ perceived sexual behaviors and limited knowledge about his serostatus, which exacerbated the risk of infant HIV transmission. Social support from families, partners and friends, were important for women who reported high adherence levels. Women in our study identified multiple barriers to PrEP adherence, including anticipated stigma and fear that people would think that PrEP was ART for HIV treatment. Women who were afraid of their partners’ or friends’ reactions managed these obstacles by disclosing their PrEP use upfront and providing information about PrEP benefits and safety during pregnancy. Women also reported concerns about safety and experiencing transient side effects. Despite these barriers, information, education from the PrEP study and empowerment of taking PrEP enabled women to overcome misgivings and continue taking and adhering PrEP. Finally, empathetic counsellors, and available PrEP services that were accessible and integrated into ANC, enabled women to start and continue taking PrEP, while ongoing adherence counselling improved women’s ability to develop strategies to adhere to daily PrEP use.

Our findings have important implications for future maternal PrEP interventions. Our participants highlighted women’s fear of partners’ risky behavior, and that partners were reluctant to test and share their serostatus. Secondary distribution of HIV self-tests, whereby female clients bring tests home to their partner(s), may be an effective way to promote HIV testing among male partners. [21-24] Further, prior research has demonstrated that partners are important barriers to women’s participation in clinical trials, especially during pregnancy,[25] and PMTCT activities more broadly [26]. Educating male partners about PrEP and providing HIV testing services may contribute to PrEP persistence and adherence for women during pregnancy and postpartum.

All women with high adherence had effectively disclosed their PrEP use to their sex partners and families, and many benefitted from social support, encouragement and adherence reminders. Similar to ART programs, disclosure and family support appears to be key to longer-term PrEP adherence and persistence. A recent study from South Africa and Zimbabwe demonstrated that stigma and disclosure were significant concerns for young women initiating PrEP.[27] Our previous analysis demonstrated the role that anticipated stigma played as a barrier to PrEP initiation and persistence in our cohort[28]. Future interventions should address stigma, especially anticipated stigma, including mentor mother or peer based programs to share how women countered stigma and negativity in their community [29].

Pregnant women may experience more side effects than other PrEP users because of concomitant morning sickness and other hormonal changes. Side effects were shown to affect PrEP continuation in pregnant women in Kenya [30] and should be addressed through education and counseling support. Women in our study were able to manage common side effects and persisted on PrEP. Some took PrEP at night when they were not prone to morning sickness. Others continued taking PrEP in postpartum period, despite abstinence during after childbirth, because they did not want to restart and experience side effects again.

Counseling about the risks, benefits, safety and adherence is essential to improving PrEP adherence, as others have demonstrated.[31-33] Women used the knowledge gained during counseling sessions to inform others about PrEP and were able to convince their partners and family to accept that PrEP was both safe and effective. Future programs should consider integrating HIV risk reduction counselling, including PrEP, by empathetic counsellors into existing ante- and postnatal care.

In South Africa, the National PrEP guidelines were recently updated to include pregnant and breastfeeding women at high risk of HIV acquisition[34]. However, this policy has not yet been translated to implementation and PrEP remains largely unavailable to pregnant/postpartum women.[35] Our hope is that the findings from our study will inform policy aimed at improving the efficiency of PrEP implementation and scale-up in South Africa.

Limitations include that the study was conducted in one urban facility in Cape Town, and may not represent rural perspectives or other regions of South Africa. We used self-reported adherence instead of objective, blood, urine or hair adherence assessments for sampling. Future studies should compare self-reports to objective tenofovir analyses for sampling in qualitative data collection.

## Conclusions

Our study identified important facilitators of optimal maternal PrEP use in pregnancy and postpartum. Individual risk, fear of HIV acquisition for self and infant, and partner sexual behaviors were important motivators to start PrEP and disclosure to family, partner and friends contributed to PrEP persistence. Maternal PrEP programs should consider how best to address barriers to PrEP use including stigma, fear around safety and side effects in the counseling around PrEP to ensure optimal PrEP use. Integration of PrEP, and adherence counseling, into existing ante- and postnatal care services will help ensure access in pregnant and postpartum women.

## Data Availability

Data available upon request from PI.

## Acknowledgements

We would like to thank our study participants, PrEP-PP study staff and the City of Cape Town Department of Health staff. We received the study drug (Truvada®) from Gilead (California, USA) and STI test kits from Cepheid (California, USA). DJD received funding from Fogarty International Center and National Institute of Health (K01TW011187), TC and LM received funding from National Institute of Mental Health (R01MH116771).

## Authorship contributions

DJD designed the study, wrote the first draft of the paper, and reviewed the data and coding for intercoder reliability

LK reviewed the study design, analysis and development of the codebook, and reviewed/revised study drafts

JMM coded the data, analyzed data in Nvivo, and revised the drafts of the paper

NT was the study interviewer, revised study transcripts, coded and analyzed the data, revised drafts of the paper

YG coded the data, analyzed data in Nvivo, and revised the drafts of the paper

NM is the study coordinator who reviewed the study implementation, data collection, coding and analysis, and reviewed/revised study drafts

KD reviewed the study design, study data and revised the final manuscript PG reviewed the study design, study data and revised the final manuscript LGB reviewed the study design, study data and revised the final manuscript

TC designed the study, reviewed the study data and revised the final manuscript LM designed the study, reviewed the study data and revised the final manuscript

